# Application of a Concise Video to Improve Patient Understanding of Tumor Genomic Testing in Community and Academic Practice Settings

**DOI:** 10.64898/2026.03.05.26347758

**Authors:** Deloris J. Veney, Lai Wei, Jaden Miller, Amanda E. Toland, Carolyn J. Presley, Heather Hampel, Tasleem J. Padamsee, Michael J. Bishop, James J Kim, Shelly R. Hovick, William J. Irvin, Leigha Senter, Daniel G. Stover

## Abstract

**Purpose:** Tumor genomic testing (TGT) is standard-of-care for most patients with advanced/metastatic cancer. Despite established guidelines, patient education prior to TGT is frequently omitted. The purpose of this study was to evaluate the impact and durability of a concise 3-4 minute video for patient education prior to TGT in community versus academic sites and across cancer types.

**Patients and Methods:** Patients undergoing standard-of-care TGT were enrolled at a tertiary academic institution in three cohorts: Cohort 1-breast cancer; Cohort 2-lung cancer; Cohort 3-other cancers. Cohort 4 consisted of patients with any cancer type similarly undergoing SOC TGT at one of three community cancer centers. Participants completed survey measures prior to video viewing (T1), immediately post-viewing (T2), and after return of TGT results (T3). Outcome measures included: 1) 10-question objective genomic knowledge/understanding (GKU); 2) 10-question video message-specific knowledge (VMSK); 3) 11-question Trust in Physician/Provider (TIPP); 4) perceptions regarding TGT.

**Results:** A total of 203 participants completed all survey timepoints. Higher baseline GKU and VMSK scores were significantly associated with higher income and greater years of education. For the primary objective, there was a significant and sustained improvement in VMSK from T1:T2:T3 (P_overall_ p<0.0001), with no significant change in GKU (p=0.41) or TIPP (p=0.73). This trend was consistent within each cohort (all p≤0.0001). Results for four VMSK questions significantly improved, including impact on treatment decisions, incidental germline findings, and insurance coverage of testing.

**Conclusions:** A concise, 3-4 minute, broadly applicable educational video administered prior to TGT significantly and sustainably improved video message-specific knowledge in diverse cancer types and in academic and community settings. This resource is publicly available at http://www.tumor-testing.com, with a goal to efficiently educate and empower patients regarding TGT while addressing guidelines within the flow of clinical practice.

## INTRODUCTION

From 2017-2021, there was an estimated 3-fold increase (9% to 30%) in the proportion of cancers for which there exists FDA-approved standard care therapy based on somatic/tumor genomic testing (TGT) result.^1^ TGT is now viewed as a necessity by most oncologists in nearly all patients with metastatic cancer^1^ and this is reflected in growing, widespread TGT use. TGT is employed in specific cancer types at diagnosis, for example, TGT guides first-line treatment of non-small-cell lung cancer.^2^ As TGT has become increasingly adopted as part of standard cancer care,^3^ ethical challenges to address variable clinical utility, incidental germline findings, and disparities around TGT options have been revealed.^4,5^ It is critical to address these challenges now as the reliance on TGT to inform treatment-related decision-making is increasing. Despite widespread use of TGT, there is documented evidence that patient pre-test education is often incomplete and varies significantly across providers.^6–9^

Application of TGT in clinical practice continues to increase and the number of mutation-directed therapies is expanding, it remains a minority of patients who receive a TGT result that leads to treatment with a mutation-matched targeted therapy. Stover et. al. previously completed a prospective trial in which 100 patients with metastatic breast cancer (MBC) underwent clinical TGT and completed five published/validated survey measures.^7,10^ This demonstrated that patients with MBC undergoing TGT *without* TGT education had less confidence in their cancer treatment,^7^ specifically reporting a significant decrease in patients’ confidence of treatment success following TGT when subsequent treatment change was not supported by the TGT results.^7^ To address this need for pre-TGT education, we completed a quality improvement cycle, identifying that the limited clinical encounter time was a major barrier to effective TGT education.^11^ In response, based on data obtained through conducting diverse patient focus groups, we developed an animated 4-minute video for patient TGT education that addresses national guidelines surrounding TGT from the American Society of Clinical Oncology (ASCO) and the American College of Medical genetics and Genomics (ACMG). Our TGT video incorporates culturally diverse images and is available in English and Spanish (viewable at http://www.tumor-testing.com). The video is publicly-available and mobile-friendly to facilitate easy access.

We hypothesized that pre-TGT education, accomplished by a concise modular video intervention adaptable to specific cancer patient populations, could provide guideline-concordant information to patients in a consistent manner. The objectives of this intervention were to communicate the benefits, limitations, and risks, including the possibility of incidental findings associated with TGT. In a preliminary analysis of 150 patients across cancer types, we demonstrated significant improvement in message-specific knowledge after viewing the video.^12^ Although our initial study suggested this concise video intervention was effective, several outstanding questions remained including durability of effect and application of the TGT educational video at academic versus community oncology practices. Differences related to TGT application have likely narrowed in recent years with more broad uptake of TGT at all care sites,^13–17^ while differences between community and academic patient populations, such as degree of education and income, have likely remained static due to barriers to care. Lastly, the impact of TGT on identification and follow-up of incidental germline findings remains unclear. Incidental germline findings are surprisingly common in the context of TGT, with 10-30% of patients undergoing tumor-germline sequencing having pathogenic or potentially pathogenic variants identified.^18,19^ Even so, although most patients with advanced cancer are interested in being notified regarding secondary germline findings from TGT, some specifically do not want testing.^4,8,20^

We present the final analyses of a prospective interventional study of a concise tumor genomic testing educational video viewed prior to completion of TGT (NCT05215769), including evaluation of patients in community and academic sites, comparison across cancer types, durability of message retention, and association of TGT with provider germline genetic testing documentation and referrals. The primary objective was to evaluate the efficacy of our educational video in diverse settings with the primary hypothesis that patient understanding of TGT (‘video message-specific knowledge’/VMSK) would be significantly increased in both academic and community settings. Secondary objectives included defining differences in baseline genetic knowledge and trust in provider between cohorts, demographic groups, and academic vs community cancer care settings. As an exploratory endpoint, we investigated the incidental germline frequency and subsequent referrals for genetic counseling.

## METHODS

### Patient Eligibility and Recruitment

The study protocol (OSU#2021C0209) was approved by the Ohio State University (OSU) Institutional Review Board (IRB) and Bon Secours Mercy IRB ceded review to the OSU as IRB of record. Participants were eligible for inclusion if they were age eighteen or older at the time of study entry with biopsy-confirmed cancer, spoke English or Spanish, and planned to undergo TGT. TGT could be from tumor tissue/cells or -liquid biopsy. Any commercial vendor of TGT or Ohio State University Molecular Pathology lab was acceptable. Eligible patients receiving care in cancer care clinics at participating sites were identified by research staff through TGT order placement, screening, or provider referral. The MBC cohort included only metastatic cancer patients while lung cancer and tumor-agnostic cohorts included patients of varied cancer stage, based on the cancer-appropriate use of TGT. A total of 203 participants were consented and completed all required surveys to be eligible for inclusion in the primary analysis (CONSORT diagram **Supp Fig 1A**) from March 2022 through December 2024 (**Supp Fig 1B**).

### Survey Instruments

#### Video message–specific knowledge (VMSK)

Our primary endpoint, video message-specific knowledge, was measured by 10 true/false statements that addressed key knowledge domains in the video intervention, with a final score reported as the number correct out of total (reported as percentage correct). Examples of statements included “The information that I get from tumor tissue genomic testing could be valuable to my children and other family members” and “I must have tumor genomic testing to continue with cancer treatment.”

#### Genomic Knowledge/Understanding (GKU)

Objective knowledge of genes/genetics was measured with 10 true/false statements based on a published genetic knowledge instrument, with a final score reported as number correct out of total (reported as percentage correct).^21^ Examples of these statements include “It is possible to see a gene with the naked eye”, and “A person’s race and ethnicity can affect how likely they are to get a disease”.

#### Trust in Physician/Provider (TIPP)

The 11-item Trust in Physician/Provider Survey^22^ uses a 5-point Likert scale. TIPP scores range from 11 to 55 with higher scores indicating greater trust in provider.^22^

#### Knowledge Sufficiency/Insufficiency surrounding TGT (KSI))

Using the Risk Information and Processing Model,^23^ participants self-assessed their perceptions of information sufficiency apposite TGT on a scale of zero to ten, where zero meant knowing nothing and ten meant knowing everything you could possibly know. Using the same scale, they then determined how much they needed to know.

#### Participant Impressions of Video

Participants responded to four statements on a scale of agreement ranging from strongly agree to strongly disagree to measure their opinion of the video. They also responded to the prompt “the amount of information in the video was” by selecting one of three options: “too little”, “theright amount”, or “too much”.

### Study Procedures

All study procedures, including informed consent, survey instruments, and video intervention were completed through a single REDCap survey via tablet provided in clinic for this use. Both English and Spanish versions of the survey instruments were provided as applicable. Participants were enrolled across four cohorts, with no mandated minimum or maximum number of patients per cohort: Cohort 1. Metastatic breast cancer (MBC); Cohort 2. Lung cancer (LC); Cohort 3. Cancer of any type (OC).; Cohort 4. Patients with any cancer type enrolled at Bon Secours Mercy Health community oncology sites (Community). Cohorts 1-3 were accrued and participated at OSU while Cohort 4 enrolled and participated at BSMH. The video viewed by each cohort differed by the modular adaptation applied to the video: OC and BSMH cohort participants viewed a 3.05-minute video; MBC cohort participants viewed the same video with the addition of a sixteen second clip indicating that, at most, four out of ten patients with MBC receive a tumor genomic test result that determines their treatment; LC cohort participants also had an additional sixteen seconds of video content indicating that three out of ten patients with LC have a tumor genomic test result that determines their treatment. All survey outcome measures were completed at timepoint 1 (T1), immediately prior to video viewing, including: demographics, GKU, VMSK, TIPP. None of the knowledge-related questions were cohort-specific. With the exception of demographics, all instruments were repeated at timepoint 2 (T2), following video viewing and prior to discussion with provider of participant’s own results. Following video-viewing, participants responded to seven questions regarding their opinions of the video that they viewed. VMSK, GKU, TIPP, and KSI were completed at timepoint 3 (T3), following return of TGT results to patient. Survey instrument utilization by time point provided **Supp Fig 1C** and full survey instrument questions provided in **Supplementary File 1.**

### Chart Review for Incidental Germline Findings

A retrospective chart review was conducted for 176 patients enrolled at OSUCCC (Cohorts 1-3) who had undergone TGT. Patients consented to chart review for five years following receipt of TGT results as part of their enrollment. European Society for Medical Oncology Precision Medicine Working Group (ESMO PMWG) guidelines were applied in three disease-focused groups: breast cancer (n=55), lung cancer (n=64), and other cancers (n=57) to determine which patients should have received germline follow up. All TGT was ordered per provider preference and included reports from several different vendors. The PDF reports from TGT analysis were located in the electronic health record (EHR) and reviewed for the presence of variants in ESMO PMWG genes attached to recommendations for germline follow up. Provider progress notes and referrals were reviewed after the date of TGT result report to determine if TGT-relevant referrals were made. In those cases where no referrals were evident, historic records were reviewed to determine if germline testing was performed pre-TGT result report.

### Statistical Analysis

Patients who completed all T1, T2, and T3 surveys were included for analyses. Demographics and baseline characteristics are summarized using descriptive statistics for the entire cohort as well as for each group (see table 1). For association of participant characteristics with baseline survey metrics, associations of VMSK, GKU, and TIPP with age of entry were explored using Spearman correlations due to uneven distributions while associations of VMSK, GKU, and TIPP with other categorical demographics were explored with non-parametric Kruskal-Wallis tests. Survey measure change over time (T1:T2:T3) was evaluated by Friedman test, a non-parametric test for repeated measures. Changes in the proportion of individuals answering specific questions correctly within VMSK between T1 and T3 were evaluated using McNemar’s test for the overall population and within each group. Only descriptive statistics were applied to qualitative endpoints: KSI and participant’s opinions of the video.

**Table 1.** Participant Characteristics.

| Variable | Level | Arm 1<br>Breast Cancer<br>(n=50) | Arm 2<br>Lung Cancer<br>(n=51) | Arm 3<br>Other Cancer<br>(n=50) | Arm 4<br>Community<br>(n=52) | Total<br>(n=203) |
| --- | --- | --- | --- | --- | --- | --- |
| Age at Diagnosis | Median<br>(min, max) | 60<br>(32, 80) | 62<br>(37, 93) | 63<br>[46, 69]<br>(18, 78) | 68<br>(29, 98) | 63<br>(18, 98) |
| Sex | Female<br>Male | 50 (100%)<br>0 (0%) | 27 (53%)<br>24 (47%) | 21 (42%)<br>29 (58%) | 33 (63%)<br>19 (37%) | 131 (65%)<br>72 (35%) |
| Education | Elementary<br>Some high school<br>High school graduate<br>Some college or<br>technical school<br>College | 1 (2%)<br>1 (2%)<br>12 (24%)<br>13 (26%)<br>23 (46%) | 0 (0%)<br>4 (8%)<br>14 (27%)<br>18 (35%)<br>15 (29%) | 1 (2%)<br>1 (2%)<br>13 (26%)<br>14 (28%)<br>21 (42%) | 0 (0%)<br>5 (10%)<br>17 (33%)<br>16 (31%)<br>14 (27%) | 2 (1%)<br>11 (5%)<br>56 (28%)<br>61 (30%)<br>73 (36%) |
| Income | < \$10,000<br>\$10,000 - \$24,999<br>\$25,000 - \$34,999<br>\$35,000 - \$49,999<br>\$50,000 - \$74,999<br>\$75,000 - \$99,999<br>\$100,000 - \$199,999<br>> \$200,000<br>Prefer not to answer | 2 (4%)<br>7 (14%)<br>6 (12%)<br>3 (6%)<br>7 (14%)<br>6 (12%)<br>12 (24%)<br>3 (6%)<br>4 (8%) | 1 (2%)<br>3 (6%)<br>2 (4%)<br>7 (14%)<br>13 (25%)<br>7 (14%)<br>10 (20%)<br>1 (2%)<br>7 (14%) | 1 (2%)<br>6 (12%)<br>4 (8%)<br>3 (6%)<br>9 (18%)<br>9 (18%)<br>8 (16%)<br>5 (10%)<br>5 (10%) | 5 (10%)<br>2 (4%)<br>5 (10%)<br>4 (8%)<br>4 (8%)<br>6 (12%)<br>2 (4%)<br>0 (0%)<br>24 (46%) | 9 (4%)<br>18 (9%)<br>17 (8%)<br>17 (8%)<br>33 (16%)<br>28 (14%)<br>32 (16%)<br>9 (4%)<br>40 (20%) |
| Marital status | Married or domestic<br>partnership<br>Single, never married<br>Divorced<br>Widowed<br>Prefer not to answer<br>Separated | 32 (64%)<br>5 (10%)<br>9 (18%)<br>2 (4%)<br>0 (0%)<br>2 (4%) | 33 (65%)<br>3 (6%)<br>6 (12%)<br>8 (16%)<br>1 (2%)<br>0 (0%) | 38 (76%)<br>7 (14%)<br>3 (6%)<br>2 (4%)<br>0 (0%)<br>0 (0%) | 24 (46%)<br>13 (25%)<br>7 (13%)<br>6 (12%)<br>0 (0%)<br>2 (4%) | 127 (63%)<br>28 (14%)<br>25 (12%)<br>18 (9%)<br>1 (0%)<br>4 (2%) |
| Race | White<br>Black or African-American<br>Hispanic, Latino, or<br>Spanish<br>Asian<br>Some other race or<br>ethnicity | 46 (92%)<br>3 (6%)<br>0 (0%)<br>1 (2%)<br>0 (0%) | 46 (90%)<br>4 (8%)<br>0 (0%)<br>0 (0%)<br>1 (2%) | 44 (88%)<br>5 (10%)<br>1 (2%)<br>0 (0%)<br>0 (0%) | 42 (81%)<br>9 (17%)<br>1 (2%)<br>0 (0%)<br>0 (0%) | 178 (88%)<br>21 (10%)<br>2 (1%)<br>1 (0%)<br>1 (0%) |
| Awareness of<br>Prior Tumor<br>Genomic Testing | I don't recall<br>No<br>Yes | 5 (10%)<br>14 (28%)<br>31 (62%) | 4 (8%)<br>32 (63%)<br>15 (29%) | 2 (4%)<br>34 (68%)<br>14 (28%) | 2 (4%)<br>41 (79%)<br>9 (17%) | 13 (6%)<br>121 (60%)<br>69 (34%) |
| Family History of<br>Cancer | I don't know<br>No<br>Yes | 2 (4%)<br>14 (28%)<br>34 (68%) | 2 (4%)<br>13 (25%)<br>36 (71%) | 4 (8%)<br>14 (28%)<br>32 (64%) | 3 (6%)<br>12 (23%)<br>37 (71%) | 11 (5%)<br>53 (26%)<br>139 (68%) |

## RESULTS

### Study Participants

Participants (n=203) who completed survey instruments at T1/T2/T3 were considered evaluable for the primary study (**Table 1).** Participant age ranged from 18 to 98 years at study entry. Most participants were female (131/203; 65%), of White race (178/203, 88%), and were married or in a domestic partnership (127/203, 63%). Education and income were relatively evenly distributed (**Table 1**). Among participants, eight distinct cancer types were represented (**Supp Fig 1C**).

### Association of Baseline Patient Characteristics with Survey Outcomes

The association of patient characteristics with baseline VMSK, GKU, and TIPP were assessed via Spearman correlations (**Table 2**). Across the entire study population there was a significant association between education and baseline/T1 VMSK (nominal p<0.0001) and GKU (nom p=0.004) as well as income and baseline/T1 GKU (nom p=0.007) and GKU (nom p=0.004). There was variation by cohort – for example, the LC and OC cohorts did not show a significant association with either education or income within that cohort (**Table 2**). There were no other significant associations between other patient characteristics and video message-specific knowledge, genomic knowledge/understanding or any patient characteristics and Trust in Physician/Provider.

**Table 2.** Association of participant characteristics with baseline survey metrics.

| Instrument | Demographic parameter | Cohort 1: Breast p-value | Cohort 2: Lung p-value | Cohort 3: Other Cancer p-value | Cohort 4: Community p-value | Overall p-value |
| --- | --- | --- | --- | --- | --- | --- |
| **T1 Video message-specific knowledge (VMSK)** | Age at entry | 0.87 | 0.24 | 0.81 | 0.43 | 0.18 |
|  | Sex | NA | 0.67 | 0.62 | 0.24 | 0.60 |
|  | Education | **0.016** | 0.12 | 0.091 | **0.007** | **<0.0001** |
|  | Income | 0.16 | 0.19 | 0.23 | 0.081 | **0.007** |
|  | Marital Status | 0.84 | 0.81 | 0.93 | 0.84 | 0.64 |
|  | Race | 0.46 | 0.69 | 0.066 | 0.054 | **0.010** |
| **T1 Genomic knowledge and understanding (GKU)** | Age at entry | 0.71 | 0.87 | 0.70 | 0.75 | 0.42 |
|  | Sex | NA | 0.50 | 0.12 | 0.82 | 0.23 |
|  | Education | 0.096 | 0.36 | 0.40 | **0.007** | **0.004** |
|  | Income | **0.036** | 0.72 | 0.66 | 0.11 | **0.004** |
|  | Marital Status | 0.79 | 0.38 | 0.37 | 0.55 | 0.63 |
|  | Race | 0.81 | 0.32 | 0.057 | 0.20 | 0.48 |
| **T1 Trust in physician provider (TIPP)** | Age at entry | 0.55 | 0.63 | 0.20 | 0.62 | 0.68 |
|  | Sex | NA | 0.13 | 0.057 | **0.011** | 0.076 |
|  | Education | 0.44 | 0.20 | 0.24 | 0.37 | 0.17 |
|  | Income | 0.23 | 0.98 | 0.22 | 0.19 | 0.072 |
|  | Marital Status | 0.64 | 0.22 | 0.40 | 0.91 | 0.34 |
|  | Race | 0.46 | 0.19 | 0.22 | 0.52 | 0.35 |

The associations of VMSK, GKU, and TIPP with age (continuous) were explored with Spearman correlations. The associations of VMSK, GKU, and TIPP with other categorical demographics were explored with Kruskal-Wallis test. Nominal p-values are reported without multiple test correction.

### Survey Measure Change and Durability Over Time

To evaluate the change over time and durability of change of the three survey measures, differences over time for each survey measure were assessed (**Table 3**). For all patients – and within each cohort – VMSK increased from T1:T2 (total population T1 VMSK mean 87.1 (median 90) to T2 VMSK mean 94.2 (median 100)) with a decline, but not back to baseline at T3 (T3 VMSK mean 91.0 (median 90)). Overall, there was a significant trend for improved and retained VMSK for the overall cohort (Friedman p<0.001) and each individual cohort (Cohort 1: Breast p<0.001; Cohort 2: Lung p<0.0001; Cohort 3: other p<0.001; Cohort 4: Community p=0.001). Interestingly, while the T1:T2 absolute improvement was the greatest in Cohort 1: Breast, the T1:T3 absolute improvement was greatest in Cohort 4: Community (**Fig 1A**), suggesting that different groups may benefit and retain the information differently. Despite significant change in VMSK over time, there was no significant change in GKU or TIPP for the overall population or any individual cohort (**Table 3**), reinforcing specific improvement seen only in VMSK appears likely to be a product of video viewing.

**Figure 1.**
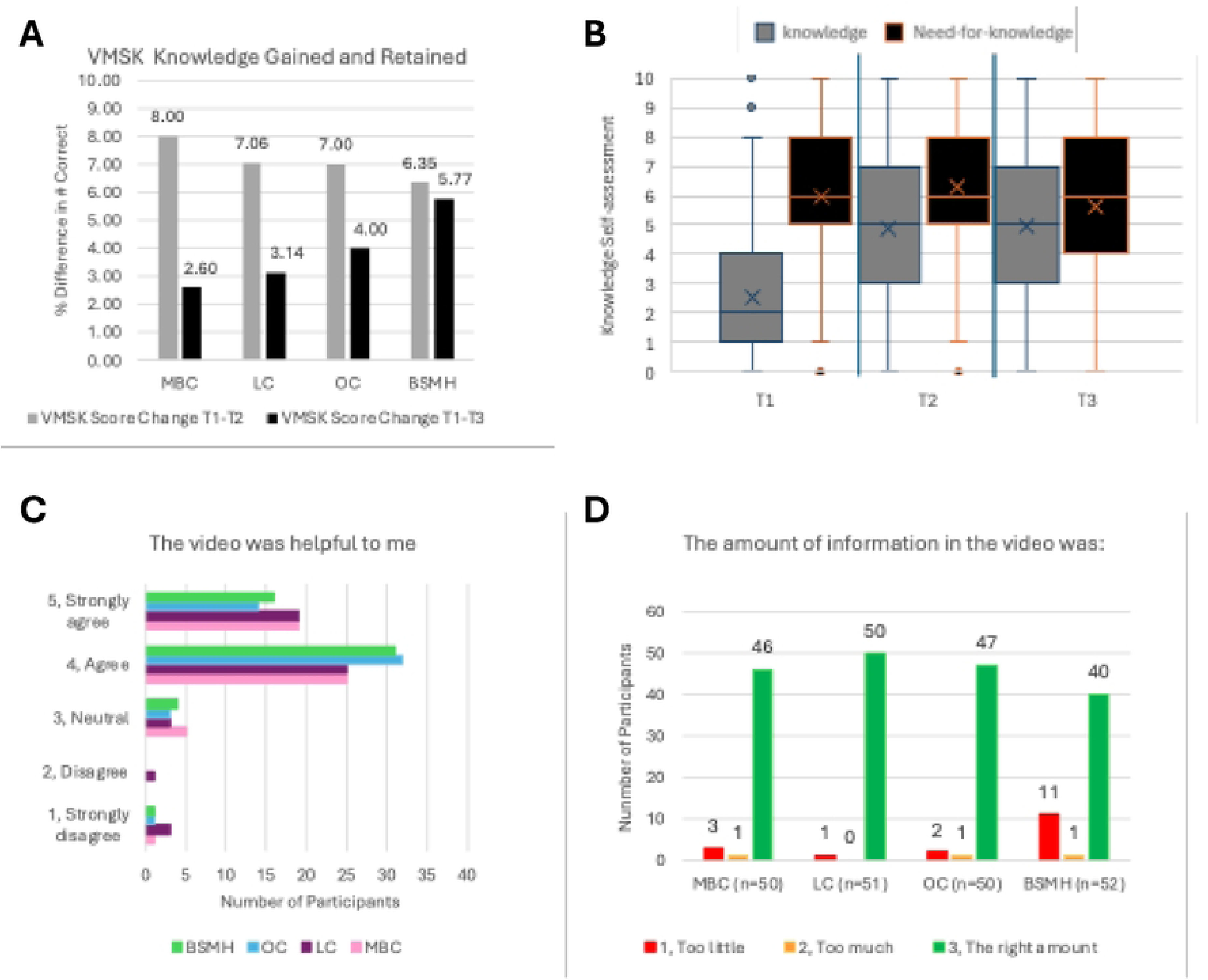
Participant retention of knowledge and perception of video content. **(A)** Video message-specific knowledge (VMSK) percent difference in number of questions correct between T1:T2 (grey) and T1:T3 (black) by cohort. (**B**) Participant self-reported TGT-related knowledge (grey) versus TGT need for knowledge, or ‘need-to-know’ (black); **(C)** Participant response to the query: “The video was helpful to me” on a 5-point Likert scale. Accrual cohort indicated by color – Cohort 1: Breast (pink), Cohort 2: Lung (purple); Cohort 3: Other (blue); Cohort 4: Community (green). (**D)**. Participant response to the query: “The amount of information in the video was:” on a 3-point Likert scale.

**Table 3.** Survey Measure Change and Durability Over Time. To evaluate the change over time of the three survey measures – video message-specific knowledge, genomic knowledge and understanding, and trust in physician/provider – differences over time were assessed using the Friedman test, a non-parametric test for repeated measures. The mean value of each measure is provided by time point and cohort. Significant (p<0.05) values indicated in bold.

| **Video Message-Specific Knowledge** |  |  |  |  |
| --- | --- | --- | --- | --- |
|  | **T1<sub>mean (median)</sub>**<br>**(%)** | **T2<sub>mean (median)</sub>**<br>**(%)** | **T3<sub>mean (median)</sub>**<br>**(%)** | **P<sub>overall</sub>** |
| ALL patients | 87.1 (90) | 94.2 (100) | 91.0 (90) | **<0.001** |
| Cohort 1 - Breast | 88.8 (90) | 96.8 (100) | 91.4 (100) | **<0.001** |
| Cohort 2 - Lung | 86.5 (90) | 93.5 (100) | 89.6 (90) | **<0.001** |
| Cohort 3 - Other | 88.0 (90) | 95.0 (100) | 92.0 (95) | **<0.001** |
| Cohort 4 - Community | 85.4 (90) | 91.7 (90) | 91.2 (100) | **0.001** |
| **Genomic Knowledge** |  |  |  |  |
|  | **T1<sub>mean (median)</sub>**<br>**(%)** | **T2<sub>mean (median)</sub>**<br>**(%)** | **T3<sub>mean (median)</sub>**<br>**(%)** | **P<sub>overall</sub>** |
| ALL patients | 84.8 (90) | 86.0 (90) | 85.8 (90) | 0.406 |
| Cohort 1 - Breast | 86.0 (90) | 85.4 (90) | 87.2 (90) | 0.676 |
| Cohort 2 - Lung | 86.7 (90) | 87.7 (90) | 84.7 (90) | 0.122 |
| Cohort 3 - Other | 86.6 (90) | 86.4 (90) | 86.6 (90) | 1 |
| Cohort 4 - Community | 80.1 (80) | 84.6 (90) | 84.6 (90) | 0.134 |
| **Provider Trust** |  |  |  |  |
|  | **T1<sub>mean (median)</sub>**<br>**(%)** | **T2<sub>mean (median)</sub>**<br>**(%)** | **T3<sub>mean (median)</sub>**<br>**(%)** | **P<sub>overall</sub>** |
| ALL patients | 46.8 (48.0) | 46.9 (48.0) | 47.2 (49.0) | 0.729 |
| Cohort 1 - Breast | 48.7 (50.5) | 48.4 (51.0) | 48.3 (50.0) | 0.66 |
| Cohort 2 - Lung | 46.4 (46.0) | 45.8 (48.0) | 47.6 (50.0) | 0.374 |
| Cohort 3 - Other | 47.3 (50.0) | 48.1 (50.0) | 47.3 (49.5) | 0.139 |
| Cohort 4 - Community | 44.7 (44.5) | 45.5 (45.0) | 45.8 (45.0) | 0.48 |

### Change Over Time of Individual Video Message-Specific Knowledge Questions

To further investigate whether a change in VMSK varied among the ten questions that comprised the VMSK instrument, as an exploratory analysis we investigated the absolute and percentage change for each individual question (**Table 4**). Only four questions demonstrated significant and retained improvement between T1:T3: 1) “I must have tumor genomic testing to continue with cancer treatment.” (p=0.028); 2) “Tumor tissue genomic results sometimes raise more questions that require more genetic testing.” (p=0.034); 3) “When my doctor has my results, they might recommend me to see a genetics specialist.” (p=0.04); and 4) “The expense of tumor genomic testing is not typically covered by health insurance.” (p=0.005). These four questions represent diverse messages including impact of TGT on treatment, potential for incidental germline findings and genetics referral, and insurance coverage of TGT. Notably, within each individual cohort few questions demonstrated significant change T1:T3 (**Supp Tables 1-4).**

**Table 4.** Change Over Time of Individual Video Message-Specific Knowledge Questions. Correct answer for each question in the video message-specific knowledge instrument, represented as number of participants (#) and percentage of total participants (%). Significant p-value (p<0.05) for change from T1:T3 indicated in bold.

| True/False Statement | T1:<br>Participants<br>correctly<br>answering |  | T2:<br>Participants<br>correctly<br>answering |  | T3:<br>Participants<br>correctly<br>answering |  | Change<br>T1:T3 |  |
| --- | --- | --- | --- | --- | --- | --- | --- | --- |
|  | # | % | # | % | # | % | # | p-value |
| We have genes in every cell of our bodies. | 191 | 94.1 | 203 | 100 | 193 | 95.1 | +2 | 0.593 |
| Tumor genomic testing might help your doctor make decisions about your cancer treatment. | 200 | 98.5 | 202 | 99.5 | 199 | 98.0 | -1 | 0.655 |
| Tumor genomic testing always determines what treatment a person will have. | 157 | 77.3 | 174 | 85.7 | 164 | 80.8 | +7 | 0.274 |
| I must have tumor genomic testing to continue with cancer treatment. | 179 | 88.2 | 187 | 92.1 | 190 | 93.6 | +11 | **0.028** |
| My doctor has other tools besides tumor genomic testing to use to choose treatments for me. | 193 | 95.1 | 199 | 98.0 | 198 | 97.5 | +5 | 0.197 |
| Tumor tissue genomic results sometimes raise more questions that require more genetic testing. | 177 | 87.3 | 194 | 95.6 | 189 | 93.1 | +12 | **0.034** |
| The information that I get from tumor tissue genomic testing could be valuable to my children and other family members. | 198 | 97.5 | 202 | 99.5 | 198 | 97.5 | 0 | 1 |
| When my doctor has my results, they might recommend me to see a genetics specialist. | 175 | 86.2 | 198 | 97.5 | 184 | 92.1 | +9 | **0.040** |
| The expense of tumor genomic testing is not typically covered by health insurance. | 114 | 56.2 | 162 | 79.8 | 139 | 68.5 | +25 | **0.005** |
| If you do not have health insurance, you cannot have tumor genomic tissue testing performed. | 185 | 91.1 | 192 | 94.6 | 191 | 94.1 | +6 | 0.180 |

### Identification of Germline Alterations via Tumor Genomic Testing

To determine the proportion of participants with potential incidental germline findings, a chart review was conducted from TGT date and forward 6 months to identify participants who would warrant follow up germline testing using the published recommendations of the European Society for Medical Oncology (ESMO) Precision Medicine Working Group guidelines.^24^ We hypothesized that TGT education will be associated with higher rates of follow-up on incidental germline findings and potentially lead to 1) genetic counseling; 2) genetic testing; and/or 3) cascade testing. Of 176 chart-reviewed patients, 18 (10%) met ESMO PMWG guidelines for germline follow-up (10/55 breast; 3/64 lung; 5/57 other) (**Table 5)**. Eight of the 18 (44%) of those with germline-relevant TGT results had previously undergone germline testing. Only 3/10 remaining patients with germline-relevant TGT results were referred for clinical genetics services or had follow up germline testing. Seven patients that were not referred for genetic follow-up had possibly germline pathogenic variants in the following 6 genes: ATM, BRCA2, CHEK2, NF1, PTEN, RET. All of these genes are associated with autosomal dominant, highly penetrant hereditary cancer risk with NCCN recommendations for gene-specific cancer screening and surveillance.

**Table 5.** Retrospective Chart Review for Germline Follow-up Testing.

| **Cancer Type** | **n** | **Met ESMO guidelines for germline follow-up** | **Had prior germline testing** | **Referred for germline follow-up** | **No germline prior testing or follow-up** |
| --- | --- | --- | --- | --- | --- |
| **Breast cancer** | 55 | 10 | 6 | 0 | 4 |
| **Lung Cancer** | 64 | 3 | 0 | 1 | 2 |
| **Other** | 57 | 5 | 2 | 2 | 1 |

### Patient Perceptions of TGT and TGT Educational Video

Participants were queried for attitudes regarding self-perceived knowledge sufficiency and knowledge ‘need-to-know’. Overall, participants related greater knowledge insufficiency at T1 with smaller gaps at T2 - and still a lesser gap at T3. A graphic representation of numerical responses is presented in **Fig 1B** to allow the reader to visualize the narrowing gap in knowledge however this is a qualitative, not quantitative measure. Similarly, most participants reported that the video was helpful (>90% of participants rating “strongly agree” or “agree”; **Fig 1C**) and most participants rated the amount of information in the video as “the right amount” (**Fig 1D).**

## DISCUSSION

Overall, this study demonstrated that a concise, 3-4 minute video improves participant video message-specific knowledge across cancer types and care delivery settings (academic vs. community). This improvement was seen across diverse topics ranging from whether TGT will impact their treatment selection to incidental germline findings to insurance coverage. Further, participant self-perceived TGT knowledge insufficiency decreased after viewing the video and the majority of participants felt that the video was helpful. This study adds to existing literature showing that implementation of a video intervention to address these gaps is feasible. Bradbury et al. (ECOG-ACRIN EAQ152) showed that a web-based genetic counseling intervention increased patient understanding but did not reduce anxiety, depression, or cancer-specific distress.^25^

The addition of a community-centered cohort was intended to increase the heterogeneity of our participant population, not only geographically, but across all directly measured demographics such as race, ethnicity, education, and income, as well as through less easily defined social and cultural parameters. Engaging patients treated in the community and collaborating with community cancer care practitioners are key components^26^ of building an educational intervention that aids in empowering patients to participate in shared decision-making across the cancer care delivery continuum.

National guidelines are unified in the need for providers review potential benefits, limitations, and risks before testing prior to TGT,^27–31^ suggesting that the goal should be 100% accuracy for the guideline-driven content within the video intervention. In this study, the significant improvement to mean 94.1% accuracy (median 100%) after video intervention represents a clinically meaningful enhancement in patient TGT knowledge. Both ASCO and ACMG also comment on the burden this places on providers as they order TGT to inform therapy selection for patients. ASCO encourages research into the delivery of this education while ACMG’s policy statement specifically puts forward innovative methods of providing this education via video or artificial intelligence. These organizations echo the importance of informed consent prior to TGT and allowing patients to opt out of receipt of germline results.^32,33^ In this study, despite published recommendations for germline follow-up of TGT results, 7/10 patients in our cohort were not referred for clinical genetic counseling nor offered germline genetic testing by their oncologist. This is consistent with, and does not demonstrate improvement over, published data on the rate of omitted genetic counseling/testing referrals.^34^ Our prior work suggests that incidental germline findings are not discussed routinely in oncology care largely due to time constraints. This lack of follow up limits the ability to detect families with the highest cancer risks and initiate risk-reducing care interventions.

Why were these instruments/endpoints selected? VMSK provides insight into the key material specifically included in the video, while GKU provides an alternative set of related data that were not specifically included in the video, thus serving as a control metric. Our prior work demonstrating diminished trust in provider choice of therapy after undergoing TGT, complements data that trust may contribute to improvements in actual health outcomes,^35^ thus trust is an important endpoint. We did not include alternative instruments addressing metrics of depression (CES-D),^36^ anxiety (BAI),^37,38^ and self-efficacy metric (CASE-cancer)^39^ based on prior work where these metrics did not change pre-/post-TGT, nor were associated with TGT-based therapy change.^10,25^ KSI allows a qualitative evaluation of patients’ perceptions of knowledge sufficiency, with expanding gaps indicative of increased information seeking and narrowing gaps indicative of satisfying patients’ desire for knowledge. This interventional video demonstrated substantial bridging of perceived health literacy gap.

In conclusion, this study is an important next step in achieving equitable TGT education for all patients. These data suggest that a concise educational video intervention improves participant message-specific knowledge across cancer types and care settings (academic and community) and that this improvement is retained in the weeks and months after video viewing. This resource is publicly available at http://www.tumor-testing.com and offers the potential to overcome provider inconsistency and time constraints by leveraging technology applications such as this video intervention.

## Acknowledgements

This work was supported by National Institutes of Health (NIH) grant 1R21CA259985 (LW, LS, SRH, HLH, AET, CJP, DGS), P30CA016058 (DGS, LW, TS, AET), 1R01 CA215151 (AET), CJP is supported by 1K76AG074923-01.

## Ethics Approval

The study protocol, OSU#2021C0209, was approved by The Ohio State University Institutional Review Board.

## Data Availability

Raw data were generated at The Ohio State University. Derived data supporting the findings of this study are available from the corresponding author, DS, on request.

